# Mapping of the country-wide prevalence of non-malarial febrile illnesses in areas with varying malaria transmission intensities in Mainland Tanzania

**DOI:** 10.1101/2024.11.28.24318159

**Authors:** Salehe S. Mandai, Angelina J. Kisambale, Daniel A. Petro, Catherine Bakari, Gervas A. Chacha, Rule Budodo, Rashid A. Madebe, Dativa Pereus, Daniel P. Challe, Ramadhani Moshi, Ruth B. Mbwambo, Grace K. Kanyankole, Sijenunu Aaron, Daniel Mbwambo, Stella Kajange, Samwel Lazaro, Ntuli Kapologwe, Celine I. Mandara, Misago D. Seth, Deus S. Ishengoma

## Abstract

Recent reports revealed a declining malaria burden, but non-malaria febrile illnesses (NMFIs) have either remained unchanged or increased. This study assessed the country-wide prevalence of NMFIs and their patterns across various malaria transmission settings in Mainland Tanzania. A cross-sectional study recruited patients aged ≥ 6 months from 86 health facilities in all 26 regions of Tanzania. All patients were tested for malaria using rapid diagnostic tests (RDTs) and the prevalence of NMFIs was determined for all patients with negative results. Logistic regression was used to determine factors associated with NMFIs. Of the 18,568 patients tested, 8,273 (44.6%) had NMFIs due to negative RDT results. Higher prevalence of NMFIs occurred in females (45.8%) than males (42.8%), adults (aged ≥ 15 years, with 50.6%) compared to under-fives (42.6%) and school children (aged 5 -< 15 years, 34.3%), and in very low (71.5%) compared to high transmission areas (33.9%). NMFIs were significantly more likely in females than in males (aOR = 1.14, 95% CI = 1.07–1.22), in very low transmission areas (aOR = 4.85, 95% CI = 4.42–5.33), adults (aOR = 1.60, 95% CI = 1.46–1.75) and under-fives (aOR = 1.60, 95% CI = 1.47–1.76). The findings show high prevalence of NMFIs overall, and higher prevalence and odds of NMFIs in females, under-fives and individuals from low and very low transmission areas. These groups should be targeted with appropriate point-of-care tests and treatment strategies.

## Introduction

Although febrile illnesses are the most common reasons for seeking medical care globally,^1,2^ diagnosis and management of these illnesses are challenging due to their diverse causes and the general nature of their symptoms which are hard to distinguish clinically.^3,4^ In malaria-endemic settings, most febrile cases are often considered to be malaria, due to the similarity of their clinical presentation leading to under-reporting and mismanagement of other causes of fevers.^5,6^ However, recent advancements in diagnostic technologies, along with intensified control efforts, have shed new light on the broader landscape of febrile illnesses, revealing a complex interplay of pathogens contributing to fevers in endemic areas.^7^ The widespread use of malaria rapid diagnostic tests (RDTs) has been pivotal in this shift. These tests swiftly and rapidly detect malaria parasites, aiding healthcare providers to make confirmatory diagnoses, but they cannot distinguish malarial from non-malarial fevers.^8^ This is because there are no point-of-care (POC) tests for other pathogens that cause non-malaria fevers^9^, and this leads to poor management of cases without malaria, as well as a missed opportunity to capture other fever-causing pathogens.^10^ With the recent decline of the malaria burden in areas which were hyper-endemic for many years, failure to detect the different causes of fevers in patients with negative RDT results is an emerging dilemma for service providers. Thus, urgent actions are required to develop POC tests for multiple pathogens and algorithms for their effective management.

In recent years, Tanzania, like many other endemic countries, has witnessed a remarkable decline in malaria transmission and disease burden in different areas.^11^ However, the decline in malaria cases does not correspond to the decrease in febrile patients in areas of different malaria endemicity. In some locations, the proportion of patients with fever but without malaria parasites has remained unchanged or increased significantly.^12^ This discrepancy highlights the increasing recognition that many febrile illnesses previously treated presumptively as malaria are actually of non-malarial origin. This is especially noticeable in areas with low malaria transmission, where a large proportion of individuals with fever test negative for malaria upon visiting health facilities. Conversely, in high transmission settings, a positive RDT result does not always indicate that malaria is the only cause of fever,^13,14^ because individuals may have developed naturally acquired immunity to malaria,^15,16^ and the presenting symptoms may result from co-infections with different pathogens. Therefore, a comprehensive, evidence-based strategy is urgently needed with a clear management algorithm for fevers not caused by or occurring concurrently with malaria infections.

While it is challenging to determine whether there is an increase in NMFIs, more work should be done to enhance our understanding of the epidemiology of these conditions and pathogens causing NMFIs. Recent reviews revealed that NMFIs are widespread in Africa and most of them are dominated by bacterial and viral aetiologies.^17–19^ A recent study in Nigeria found that 37.0% of febrile cases were not caused by malaria. Among these cases, 54.7% were due to viral infections, while bacterial infections caused 32.1% of the infections.^20^ Similarly, a study in Senegal identified other causes of fever in 23.0% of febrile cases.^7^ In another study conducted in Guinea-Bissau, respiratory viruses were the most common pathogens, detected in 58.9% of non-malarial febrile cases, followed by bacteria which were found in 22.6% of the cases.^21^

In contrast to malaria, other causes of febrile illnesses are frequently unnoticed in many regions of Sub-Saharan Africa due to the lack of comprehensive guidelines for managing febrile cases when malaria is not the cause.^6,22^ This neglect stems from limited human and financial resources,^23^ and a lack of understanding and insufficient appreciation of the public health burden posed by NMFIs.^24^ The limited awareness is further compounded by inadequate training and diagnostic capacity, which hinders the ability of healthcare workers to correctly identify and appropriately manage NMFIs.^25–27^ In many regions, diagnostic tools for these illnesses are either unavailable or underutilised due to logistical challenges, leading to presumptive treatment. As a result, authorities are compelled to prioritise diseases deemed more threatening to public health, while NMFIs are overlooked due to insufficient evidence of their true burden.^28^ Subsequently, the lack of evidence-based information on the burden of these illnesses further complicates the situation, leading to disregard for the potential threat of NMFIs.^29^ This cycle of neglect continues as limited data on NMFIs hampers the development of effective policies and guidelines, and diminishing incentives to invest in diagnostic capacity. In addition, the decline of malaria is found to be associated with an increase in antibiotic prescriptions,^30,31^ highlighting the potential presumptive treatment of NMFI cases with antibiotics, which is inappropriate and could result in antibiotic resistance, a growing global concern.^32,33^

Healthcare workers in Tanzania, like in other countries, face difficulties when evaluating febrile patients, especially those with negative malaria RDT results, due to the absence of a clear evidence-based management guideline for NMFIs. This situation underscores the need for a comprehensive understanding of the country’s epidemiology and the burden of these illnesses. Although previous studies conducted in Tanzania focused on identifying aetiological agents of NMFIs at selected sites, the extensive burden and distribution patterns of NMFIs remain poorly understood. This study utilised the largest data generated by the ongoing project on molecular surveillance of malaria in Mainland Tanzania (MSMT),^34–36^ and aimed to map the country-wide prevalence and distribution patterns of NMFIs across various regions located in different malaria transmission strata. The findings from this study will inform future research and healthcare planning, and potentially improve the diagnosis and management of febrile illnesses beyond malaria, in areas of varying endemicity in the context of the ongoing epidemiological transition of malaria transmission intensities.

## Materials and methods

### Study design

This was a health facility-based cross-sectional study which was conducted in all 26 regions of Mainland Tanzania from January to August 2023. The survey was conducted under the MSMT project which focuses on mapping parasite populations, and drug and diagnostic resistance (by detecting the status of histidine-rich protein 2/3 *(hrp2/3)* gene deletions) as described elsewhere.^34–38^

### Selection of study sites in all 26 regions of Mainland Tanzania

This was the first-ever survey of the MSMT to cover all 26 regions of Mainland Tanzania and it was conducted at HFs compared to national community surveys such as school parasitological surveys,^39^ malaria indicator surveys^40–42^ and demographic and health surveys.^11^ The regions which were covered included Arusha, Dar es Salaam, Dodoma, Geita, Iringa, Kagera, Katavi, Kigoma, Kilimanjaro, Lindi, Manyara, Mara, Mbeya, Morogoro, Mtwara, Mwanza, Njombe, Pwani, Rukwa, Ruvuma, Shinyanga, Simiyu, Singida, Songwe, Tabora, and Tanga (**Figure 1**). Based on the 2022 malaria stratification by the National Malaria Control Programme, the regions were categorised into four strata high (10 regions), moderate (8 regions), low (5 regions) and very low malaria transmission intensities (3 regions) (NMCP, unpublished data). In the high transmission stratum, the regions included Geita, Katavi, Lindi, Mara, Mtwara, Pwani, Ruvuma, Shinyanga, Tabora and Tanga. The regions in the moderate stratum were Kagera, Kigoma, Mbeya, Morogoro, Mwanza, Rukwa, Simiyu and Songwe while the low transmission stratum had Dar es Salaam, Dodoma, Iringa, Njombe and Singida, and the very low transmission stratum had Arusha, Kilimanjaro, and Manyara region (**Figure 1**).

**Figure 1:**
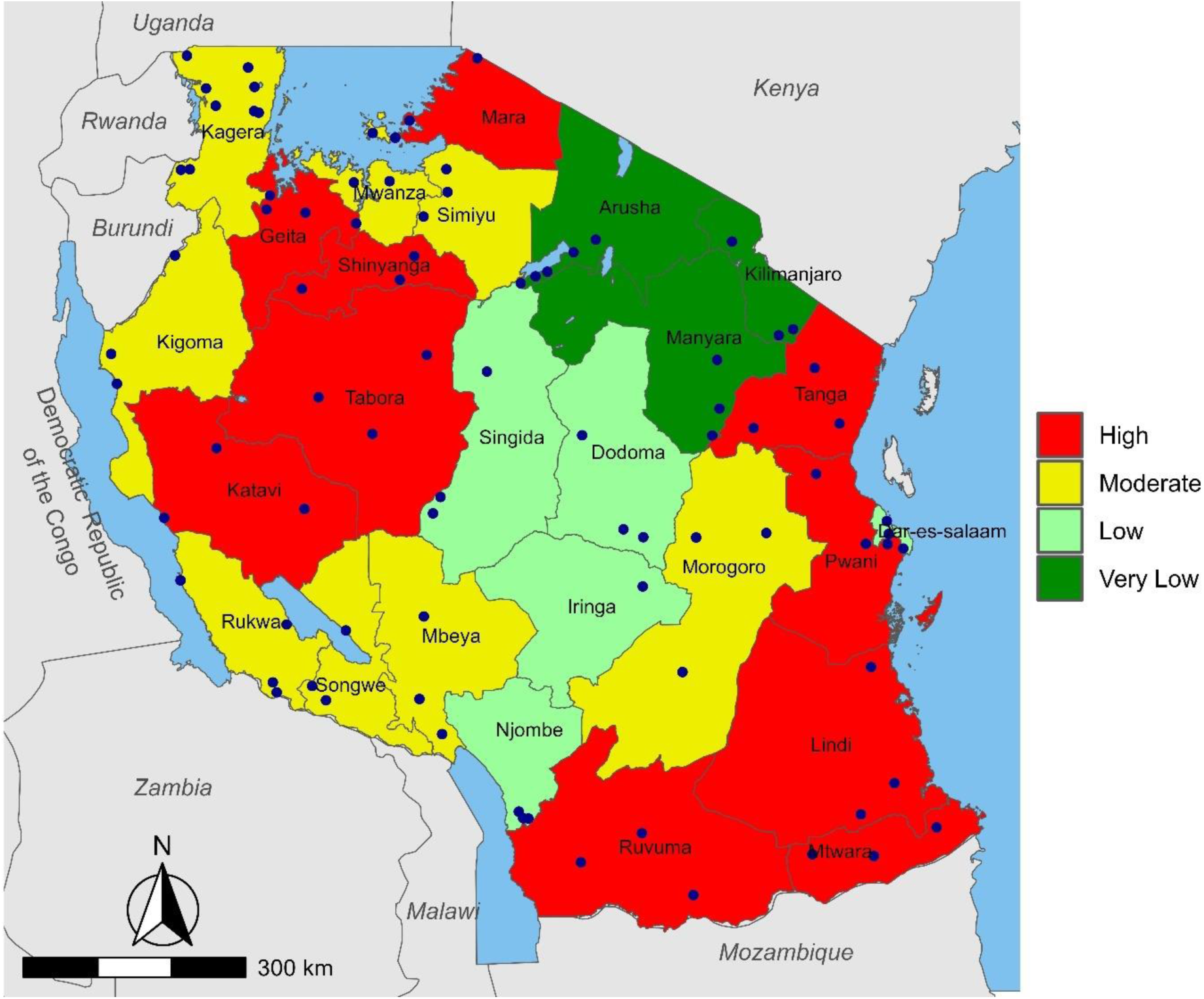
Map showing the 26 regions of Mainland Tanzania, malaria transmission strata and the health facilities (blue dots) where the data were collected.

In all 26 regions, 86 health facilities (HFs) were selected and involved in this study. In the 10 regions (Dar es Salaam, Dodoma, Kagera, Kilimanjaro, Manyara, Mara, Mtwara, Njombe, Songwe and Tanga) which were covered by the MSMT project in 2021, the selection was made within the 10 HFs which were sampled in 2021. Out of these 10 HFs, three facilities were purposively selected for the 2023 survey, with a preference to the facilities which were also covered in 2022. The process of selecting the HFs which were covered by the MSMT surveys in 2021 has been previously described.^34,38^

In each of the remaining 16 regions, three HFs from preferably three administrative councils were purposively selected to represent the diverse parts of the region and to ensure the region was appropriately represented. The selection process was done with the support of the teams from the regional and district medical officers. Initially, the data of each region was assembled from the District Health Information System 2 (DHIS2) and assessed to determine councils and HFs which reported a minimum of 50 malaria patients per month between December 2022 and April 2023. These were considered to have the capacity to recruit the required number of patients (which was 100 patients) within three months. After selecting the councils and HFs with a sufficient number of patients, the target councils were selected to ensure their distribution forms a triangular spread across the region. The selection of HFs in the region/council also considered the location of facilities which were selected in the neighbouring region to avoid over-sampling in a similar geographic area in case selection was already done in other regions close to it. Thereafter, the HFs were selected by considering the availability of sufficient staff to ensure participation in the project did not impact the provision of health services to their clients. The HFs were finally selected if they were reachable by road especially during the rainy season because the study was done before, during and after the long rainy season which normally occurs between March and June.

In all regions, three councils (with one facility in each council) were involved in the survey, except for Kagera (7 Councils and 10 HFs), Kilimanjaro, Manyara and Singida region (two councils/region), and Iringa and Arusha (one council in each region). The low number of councils in Arusha, Iringa, Kilimanjaro, Manyara and Singida was due to low transmission intensities of malaria. Therefore, very few HFs in these regions were capable of recruiting a minimum sample of 100 patients with RDT positive results which was an important requirement for a site to be selected by the MSMT project. The inclusion of more councils and HFs in Kagera was due to intensive surveillance which is currently going on in the region following the detection and confirmation of artemisinin partial resistance (ART-R).^43–45^

### Study population and participant enrolment

The study recruited outpatients aged 6 months and above who presented at the HFs and were suspected to have uncomplicated malaria based on a history of fever in the past 48 hours or fever at presentation (with axillary temperature ≥37.0℃). The inclusion criteria included febrile illnesses which were suspected to be uncomplicated malaria based on a history of fever within 48 hours before the survey or fever at presentation, age ≥6 months, and residence in an area covered by selected HF under the study. Before recruitment, health facility staff who were involved in the survey provided information about the study to all outpatients and asked eligible patients for their consent to participate. All patients who met the inclusion criteria were invited to meet the study staff who obtained informed consent and finalised the enrolment process. Consenting was done as previously described^38^ and patients who did not consent to participate or were not residents of the study area were excluded. In addition, patients with life-threatening illnesses such as severe malaria and other conditions that required immediate care were excluded from the study.

### Data collection

For this study, all febrile patients who were recruited at the HFs while seeking care and tested RDT-negative for malaria were considered to have NMFIs. For all eligible patients, demographic, anthropometric, and parasitological data were collected after consenting as shown in Figure 2. The data were collected by trained staff who participated in the 2021 or 2022 surveys of the MSMT project in the 10 regions of Dar es Salaam, Dodoma, Kagera, Kilimanjaro, Manyara, Mara, Mtwara, Njombe, Songwe and Tabora. All the staff received a refresher training for 2-3 days before initiation of the survey. In the 16 regions which took part in the MSMT project for the first time, all staff members from the selected HFs were trained by the project team. The training team included an experienced clinician and a laboratory expert. The training was done for 3 days and covered the study protocol, study documents such as informed consent forms (ICFs), case report forms (CRFs) and study logs. It also covered the basics of good clinical practices (GCP), good clinical laboratory practices (GCLP) and standard operating procedures (SOPs) of the project. After the training, HF staff conducted the enrolment of patients which included providing study information, obtaining informed consent, performing clinical assessment of all eligible patients, filling the ICFs and CRFs, performing a laboratory test and collecting samples as dried blood spots on filter papers (DBS).

**Figure 2:**
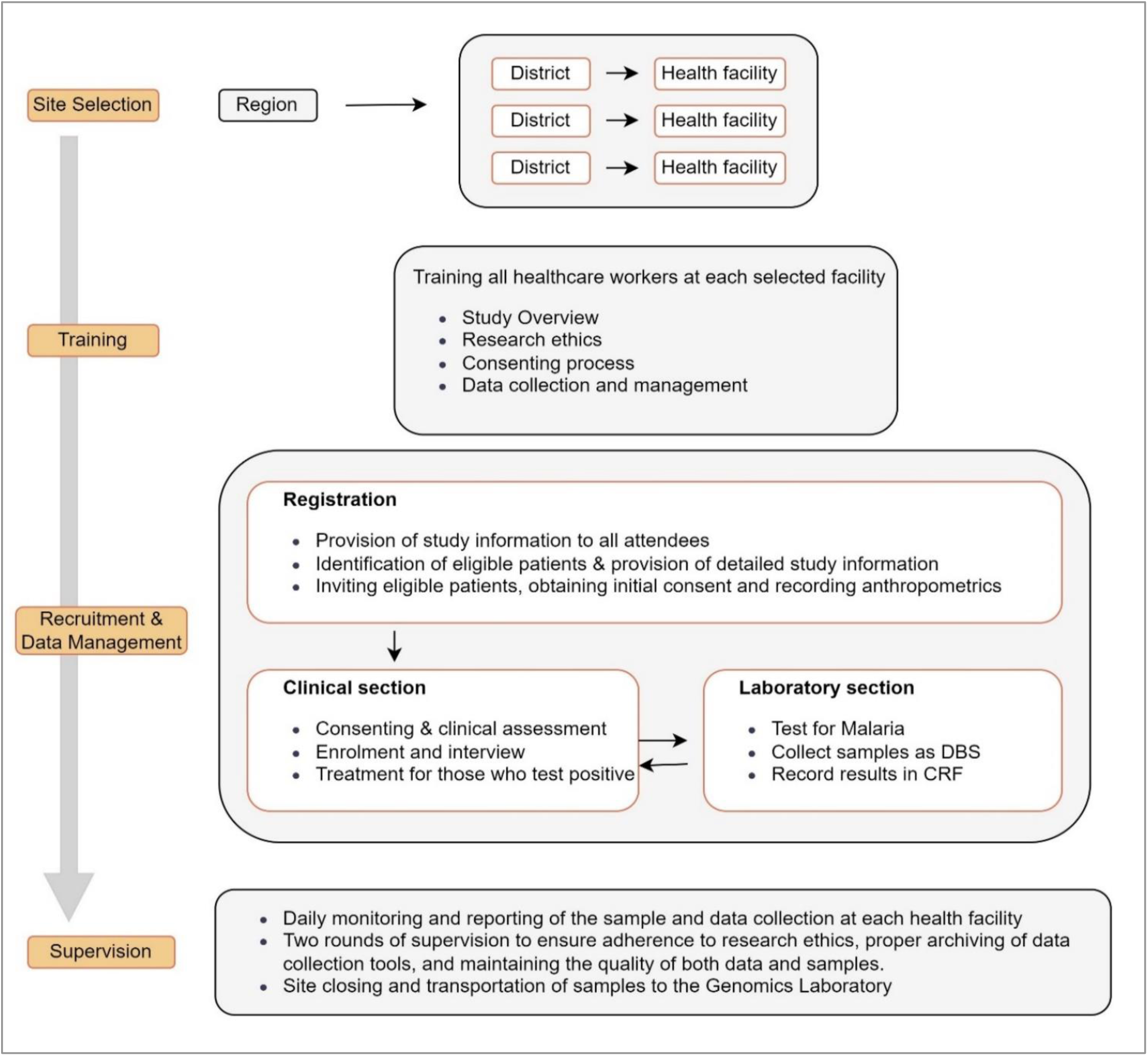
Schematic presentation of the selection of districts, health facilities and data collection process.

The data collected included basic demographics (such as residence, age and weight), axillary temperature, weight, history of fever in the past 48 hours, history of travel in the past 30 days and history of antimalarial medication in the past 14 days. Testing for malaria was done for all enrolled patients using RDTs and three brands were used in different regions/councils. All RDTs were Pf/Pan malaria and included Bioline™ Malaria Ag P.f/Pan (Abbott Diagnostics Korea, Inc., Gyeonggi-do, Korea), First Response (Premier Medical Corporation Pvt. Ltd., Gujarat, India), and SD Bioline (Standard Diagnostic Inc, Suwon, Korea). The testing process was done following routine procedures for outpatients per the manufacturer’s instructions. All the information, including RDT results, was recorded on CRFs. Each HF provided a daily report to the project team through a WhatsApp group that was set up for all HFs in the region. The report presented the number of patients screened and enrolled daily and the number of patients with positive results who were recruited. The report also presented the status and progress of the survey and the challenges encountered if any. In case the staff at the HF had critical problems such as a limited understanding of study procedures which limited the progress of the survey, they were re-trained virtually or visited by the project team.

Quality control processes included daily and weekly reviews of the data collected at each HF by the local team itself or with the support of the MSMT team (virtually). This was done to ensure the study documents (ICF, CRF and logs) were appropriately and correctly filled, and the documents were properly stored. To further ensure high-quality data and samples were collected, each HF was supervised by the MSMT team at least twice after the training. The supervision was done before the HF team collected half of the target samples and during the closure of the survey, once all the samples were collected. During supervision, the project team assessed the progress of the survey to ensure compliance with the protocol, GCP/GCLP and SOPs. The study team assessed all study documents and samples, identifying and resolving any discrepancies. The storage of samples and data was also evaluated, along with stock levels, to ensure sufficient materials were available at the facility. In case poor samples or data were collected, they were rejected, and the team was retrained and advised to restart the sample and data collection process.

### Data management and statistical analysis

Data were collected using paper questionnaires (the CRF) and double-entered into a Microsoft® Access® LTSC MSO (Version 2403) database. Data cleaning involved exporting raw data to the Microsoft® Excel® LTSC MSO (Version 2403), removing duplicate entries, and handling missing values. Cleaned data were processed and analysed using STATA Software version 13 (StataCorp LP, College Station, TX, USA) and R Software version 4.3.2 (The R Foundation for Statistical Computing, Vienna, Austria). A descriptive analysis including median, frequencies, and proportions was done to understand the data’s basic patterns and distributions. Inferential statistics involved chi-square and logistic regression to determine the statistical association between NMFIs and risk variables such as sex, age group and transmission strata of malaria. All variables with p-value < 0.25 in the univariate logistic regression were included in the multivariate regression model to understand their individual and combined effects while controlling for potential confounding factors. The association between variables were presented as crude (cORs) and adjusted odds ratios (aORs), with 95% confidence intervals (CIs), and a p-value of ≤ 0.05 was considered significant.

## Results

### Baseline characteristics of study patients

A total of 18,568 patients were recruited from 86 HFs in all 26 regions of Mainland Tanzania. The majority (57.8%, n = 10,724/18,568) of the patients were females while 42.2%, (n = 7,844/18,568) were males, and this pattern was observed in all regions except in Iringa region where 50.5% (n = 48/95) were male and 49.5% (n = 47/95) were female. The median age (interquartile range) of the patients was 9.0 (3 – 26) years, with a high number of patients aged ≥ 15 years old (41.8%, n = 7,762/18,568), followed by under-fives (41.4%, n = 7,684/18,568), and school children (aged 5 - < 15 years) (16.8%, n = 3,122/18,568). The average number of patients per region was 714, but Dar es Salaam, Kagera, Kilimanjaro, and Tabora regions contributed over 30.0% of the study patients with ≥ 1,004 patients per region. All the remaining regions had over 400 patients except Iringa region which had 95 patients only (recruitment was done by one HF due to a lack of facilities with a sufficient number of patients). Based on the strata of malaria transmission intensities, 17.8% (n = 3,311/18,568) of the patients were from very low, 16.0% (n = 2,979/18,568) from low, 31.8% (n = 5,906/18,568) from moderate and 34.3% (n = 6,372/18,568) were from regions located within the high transmission strata (**Table 1**).

**Table 1:** Demographic characteristics of the study patients.

| Region/transmission strata | N | Male | Female | < 5 years | 5 -<15 years | ≥ 15 years |
| --- | --- | --- | --- | --- | --- | --- |
|  |  | n (%) | n (%) | n (%) | n (%) | n (%) |
| Very low |  |  |  |  |  |  |
| Arusha | 561 | 259 (46.2) | 302 (53.8) | 198 (35.3) | 94 (16.8) | 269 (47.9) |
| Kilimanjaro | 2,154 | 882 (40.9) | 1,272 (59.1) | 308 (14.3) | 238 (11.1) | 1,608 (74.6) |
| Manyara | 596 | 261 (43.8) | 335 (56.2) | 165 (27.7) | 113 (18.9) | 318 (53.4) |
| Sub-Total | 3,311 | 1,402 (42.3) | 1,909 (57.7) | 671 (20.3) | 445 (13.4) | 2,195 (66.3) |
| Low |  |  |  |  |  |  |
| Dar-es-salaam | 1,117 | 558 (50.0) | 559 (50.0) | 340 (30.4) | 161 (14.4) | 616 (55.2) |
| Dodoma | 841 | 359 (42.7) | 482 (57.3) | 320 (38.1) | 138 (19.4) | 383 (44.5) |
| Iringa | 95 | 48 (50.5) | 47 (49.5) | 44 (46.3) | 26 (27.4) | 25 (26.3) |
| Njombe | 402 | 177 (44.0) | 225 (56.0) | 102 (25.4) | 104 (25.9) | 196 (48.7) |
| Singida | 524 | 226 (43.1) | 298 (56.9) | 341 (65.1) | 51 (9.7) | 132 (25.2) |
| Sub-Total | 2,979 | 1,368 (45.9) | 1,611 (54.1) | 1,147(38.5) | 480 (16.1) | 1,352 (45.4) |
| Moderate |  |  |  |  |  |  |
| Kagera | 2,352 | 992 (42.2) | 1,360 (57.8) | 1,333 (56.7) | 328 (13.9) | 691 (29.4) |
| Kigoma | 460 | 199 (43.3) | 261 (56.7) | 240 (52.2) | 60 (13.0) | 160 (34.8) |
| Mbeya | 560 | 228 (40.7) | 332 (59.3) | 208 (37.1) | 82 (14.6) | 270 (48.2) |
| Morogoro | 476 | 161 (33.8) | 315 (66.2) | 184 (38.7) | 98 (20.6) | 194 (40.7) |
| Mwanza | 455 | 204 (44.8) | 251 (55.2) | 222 (48.8) | 106 (23.3) | 127 (27.9) |
| Rukwa | 662 | 265 (40.0) | 397 (60.0) | 267 (40.3) | 150 (26.7) | 245 (37.0) |
| Simiyu | 477 | 171 (35.8) | 306 (64.2) | 250 (52.4) | 74 (15.5) | 153 (32.1) |
| Songwe | 464 | 195 (42.0) | 269 (58.0) | 156 (33.6) | 118 (25.4) | 190 (40.0) |
| Sub-Total | 5,906 | 2,215 (40.9) | 3,491 (59.1) | 2,860 (48.4) | 1,016 (17.2) | 2,030 (34.4) |
| High |  |  |  |  |  |  |
| Geita | 461 | 200 (43.4) | 261 (56.6) | 245 (53.2) | 95 (20.6) | 121 (26.3) |
| Katavi | 578 | 201 (34.8) | 377 (65.2) | 201 (34.8) | 107 (18.5) | 270 (46.7) |
| Lindi | 426 | 175 (41.1) | 251 (58.9) | 269 (63.2) | 49 (11.5) | 108 (25.3) |
| Mara | 907 | 382 (42.1) | 525 (57.9) | 246 (27.1) | 270 (29.8) | 391 (43.1) |
| Mtwara | 820 | 348 (42.1) | 472 (57.6) | 522 (63.7) | 78 (9.5) | 220 (26.8) |
| Pwani | 667 | 287 (43.0) | 380 (57.0) | 315 (47.2) | 121 (18.1) | 231 (34.6) |
| Ruvuma | 526 | 246 (46.8) | 280 (53.2) | 245 (46.6) | 122 (23.2) | 159 (30.2) |
| Shinyanga | 430 | 180 (41.9) | 250 (58.1) | 219 (50.9) | 97 (22.6) | 114 (26.5) |
| Tabora | 1,004 | 435 (43.3) | 569 (56.7) | 605 (60.2) | 1015 (10.1) | 273 (29.7) |
| Tanga | 553 | 205 (37.1) | 348 (62.9) | 139 (25.1) | 141 (25.5) | 273 (49.4) |
| Sub-Total | 6,372 | 2,659 (41.7) | 3,713 (58.3) | 3,006 (47.2) | 1,181 (18.5) | 2,185 (34.3) |
| Grand total | 18,568 | 7,844 (42.2) | 10,724 (57.8) | 7,684 (41.4) | 3,122 (16.8) | 7,762 (41.8) |

### Prevalence of non-malarial febrile illnesses

Of the 18,568 patients tested, 8,273 (44.6%) had a negative test for malaria by RDT and were considered to have NMFIs (**Table 2**). Of all regions, Dar-es-salaam and Kilimanjaro had the highest prevalence of NMFIs (70.9%, n = 792/1,117 and 85.6%, n = 1,844/2,154, respectively) while Njombe had the lowest prevalence with 15.4% (n = 62/402) (**Figure 3A**). The prevalence of NMFIs was significantly higher in females (45.8%, n = 4,913/10,724) than in males (42.8%, n = 3,360/7,844). This pattern was observed in all regions except in Iringa where the prevalence was higher in males (64.6%, n = 31/48) than in females (57.5%, n = 27/47). The proportion of NMFI was significantly higher in adults (50.6%, n = 3,927/7,762) compared to under-fives and school children (42.6%, n = 3,276/7,684 and 34.3%, n = 1,070/3,122, respectively). In the male population, the high prevalence was observed in Kilimanjaro (80.4%, n = 709/882), Dar-es-Salaam (68.1%, n = 380/558), and Iringa (64.6%, n = 31/48), while Kigoma, Shinyanga, Lindi and Njombe regions had lower prevalence with 19.6% (n = 39/199), 19.4% (n = 35/180) 19.4% (n = 34/173) and 11.3% (n = 20/177), respectively. Among females, Kilimanjaro (89.2%, n = 1,135/1,272), Dar-es-Salaam (73.7%, n = 412/559), and Dodoma (64.7%, n = 312/482) also had a high prevalence of NMFIs, reflecting the trend observed in males. Regions such as Lindi (24.7%, n = 62/251), Mtwara (24.6%, n = 116/472), Shinyanga (19.6%, n = 49/250), and Njombe (18.7%, n = 42/225) had lower prevalence in the female population (**Figure 3B and 3C**).

**Figure 3:**
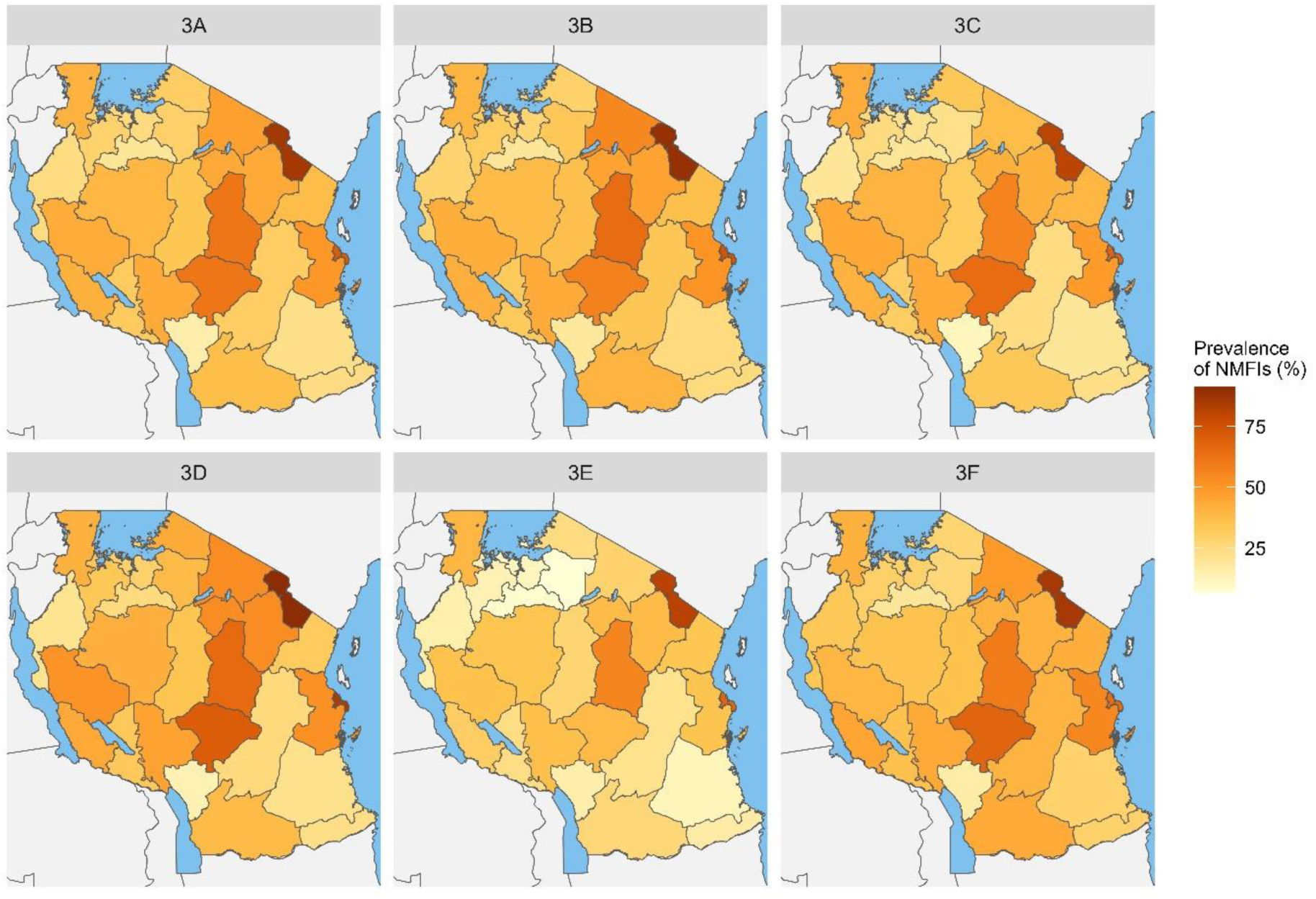
Spatial distribution of non-malarial febrile illness prevalence in six demographic groups: (3A) all patients, (3B) females, (3C) males, (3D) under-fives, (3E) school children, and (3F) adults. Each panel highlights the variation in NMFI prevalence across the study region for the specified subgroup.

**Table 2:** Prevalence of NMFI by sex, age group and malaria transmission strata.

| Variable | Febrile patients tested for malaria | Patients with negative malaria tests | Proportion of NMFI (%) | P-value |
| --- | --- | --- | --- | --- |
| Overall prevalence | 18,568 | 8,273 | 44.6 |  |
| Sex |  |  |  |  |
| Male | 7,844 | 3,360 | 42.8 | < 0.001 |
| Female | 10,724 | 4,913 | 45.8 | < 0.001 |
| Age group |  |  |  |  |
| < 5 years | 7,684 | 3,276 | 42.6 | < 0.001 |
| 5 -< 15 years | 3,122 | 1,070 | 34.3 | < 0.001 |
| ≥ 15 years | 7,762 | 3,927 | 50.6 | < 0.001 |
| Transmission strata |  |  |  |  |
| Very Low | 3,311 | 2,368 | 71.5 | < 0.001 |
| Low | 2,979 | 1,605 | 53.9 | < 0.001 |
| Moderate | 5,906 | 2,140 | 36.2 | < 0.001 |
| High | 6,372 | 2,160 | 33.9 | < 0.001 |

In under-fives, the regions of Kilimanjaro (91.2%, n = 281/308), Dar-es-Salaam (84.1%, n = 286/340), Iringa (70.5%, n = 31/44) and Dodoma (65.3%, n = 209/320) had a higher prevalence of NMFIs, indicating a significant burden of NMFIs among young children. On the contrary, regions like Njombe (13.7%, n = 14/102), Kigoma (20.8%, n = 50/240), and Lindi (21.9%, n = 59/269) had a lower prevalence of NMFIs in under-fives. Among school children, Kilimanjaro (81.1%, n =193/238) and Dar-es-Salaam (67.1%, n = 108/161) had a higher prevalence of NMFIs, while Simiyu (6.8%, n = 5/74), Shinyanga (11.3%, n = 8/97), and Mwanza (11.3%, n = 12/106) had lower prevalence in this age group. In the adult population, the highest prevalence was observed in Kilimanjaro region (85.2%, n = 1,370/1,608), while Njombe (16.3%, n = 32/196) had the lowest prevalence (**Figure 3D-F** and **Supplementary Table 1**). The proportion of NMFI was significantly lower in the high transmission stratum (33.9%) compared to the very low transmission stratum (71.5%) as shown in **Figure 4**.

**Figure 4:**
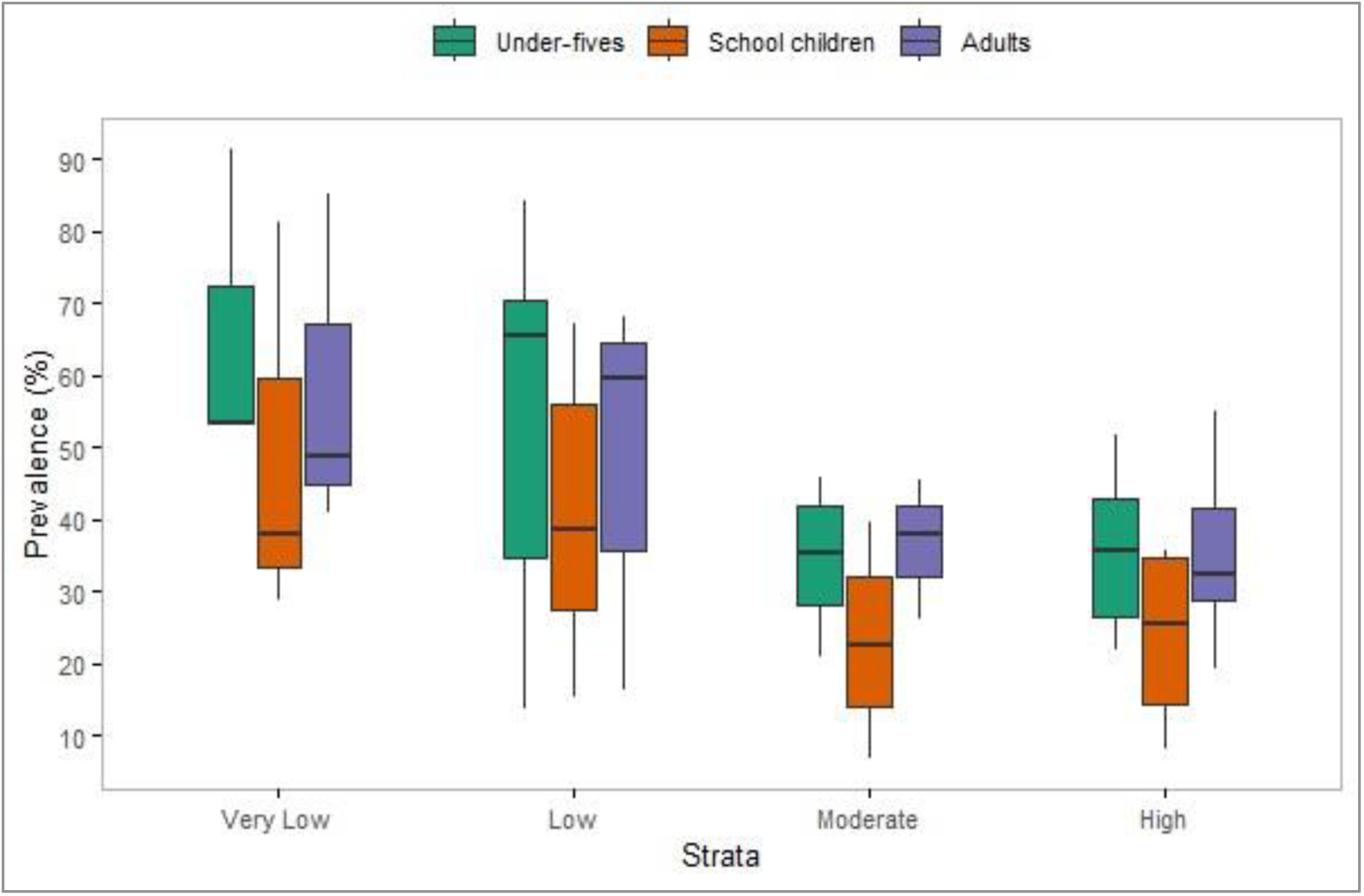
Boxplots showing the prevalence of NMFIs among patients of different age groups in the strata with different malaria transmission intensities.

### Predictors/risk factors of non-malarial illnesses

In the univariate analysis, sex, age group, and malaria transmission strata were significantly associated with the occurrence of NMFIs (p < 0.001). When all these factors were simultaneously included in a multivariate model, the odds of getting NMFI were significantly higher in females compared to males (adjusted odds ratio [aOR] = 1.14, 95% CI = 1.07 - 1.22, p < 0.001). Compared to school children, the odds of getting NMFI were significantly higher in under-fives (aOR = 1.60, 95% CI = 1.47 - 1.76, p < 0.001) and adults (aOR = 1.60, 95% CI = 1.46 - 1.75, p < 0.001). Moreover, the odds of having NMFIs were significantly higher in individuals living in settings with moderate (aOR = 1.10, 95% CI = 1.02 - 1.19, p < 0.011), low (aOR = 2.28, 95% CI = 2.09 - 2.50, p < 0.001) and very low transmission intensities of malaria (aOR = 4.85, 95% CI = 4.42 - 5.33, p < 0.001); compared to those living in the settings transmission with higher transmission intensities. The odds increased significantly across the strata, with the highest odds in the stratum with very low transmission of malaria transmission intensities as shown in **Table 3**.

**Table 3:**
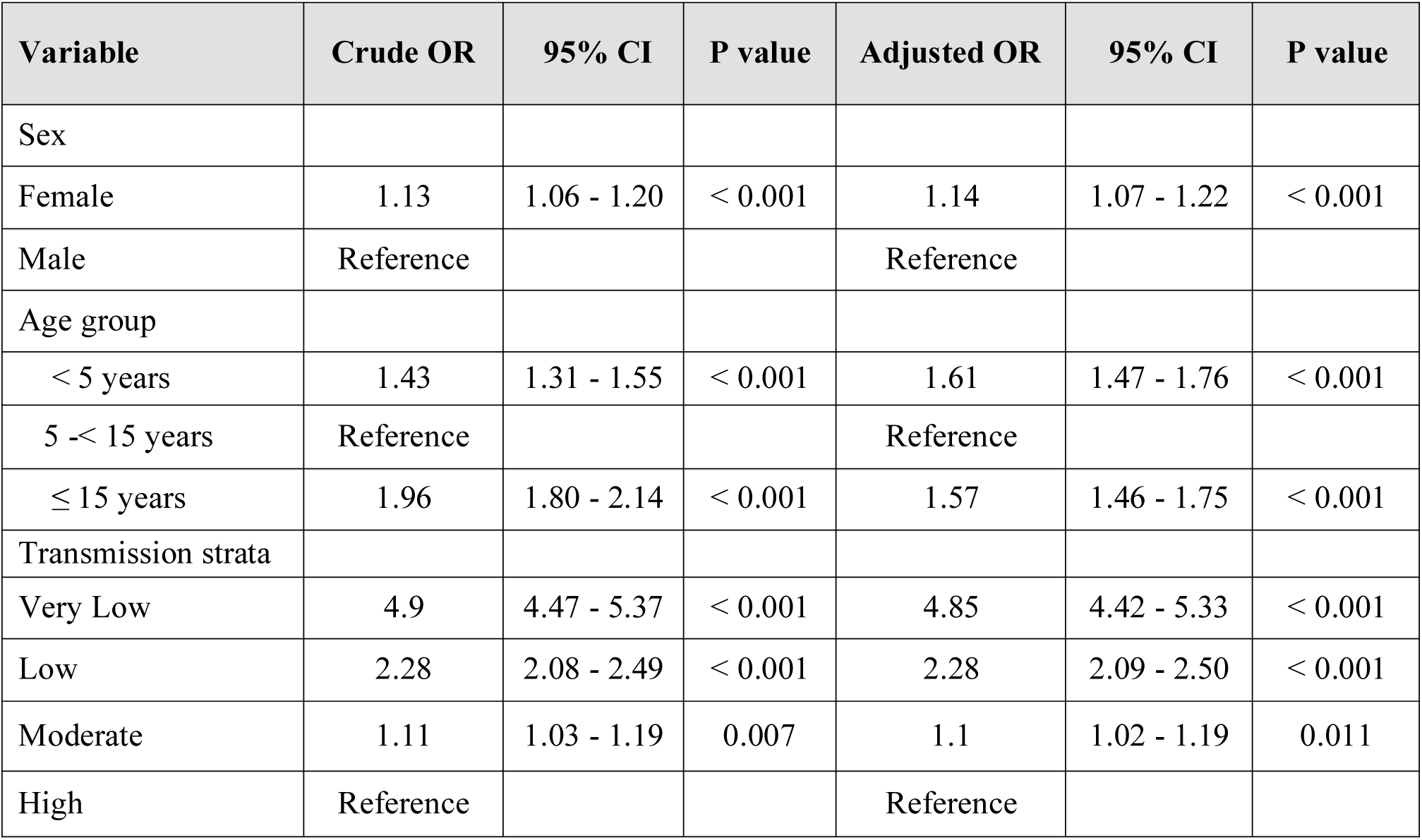
Demographic factors associated with NMFIs.

## DISCUSSION

This was the first health facility-based cross-sectional study involving symptomatic patients suspected of uncomplicated malaria to be conducted by the MSMT project covering all 26 regions of Mainland Tanzania. Unlike community surveys which are routinely implemented either in school children,^39^ or under-fives,^11,40–42^ it provides the largest scan of malaria in HF settings in the entire country. The data have also been utilised to assess the prevalence and patterns of NMFIs in Tanzania and provide useful information for guiding policy and decisions for controlling different pathogens associated with NMFIs. The study found that more than 40.0% of the febrile patients had NMFI and there was a high prevalence of NMFIs with spatial heterogeneity across different regions with varying malaria transmission intensities in Mainland Tanzania. Occurrence and the odds of NMFIs were higher in females than males, and in under-fives compared to older age groups. School children had the lowest prevalence and correspondingly, the odds of getting NMFIs. NMFIs varied inversely to malaria transmission intensities, with the lowest prevalence in the regions located in the stratum with high transmission intensities and the highest prevalence was in the stratum with very low transmission intensities of malaria. The findings provide country-wide scans of NMFIs with varying prevalence which suggest that more studies are urgently needed to further characterise the burden of NMFI, and develop POC tests and the guidelines/recommendations for managing these illnesses alongside malaria.

The findings showed that the prevalence of NMFIs in the different regions of Mainland Tanzania increased with decreasing transmission intensities of malaria. This may or may not represent a direct association between the occurrence of NMFIs and the transmission intensity of malaria. In areas with high malaria transmission, the rate of individuals harbouring malaria parasites is high compared to areas with low transmission. However, most of the people in high transmission tend to have high acquired immunity due to high rates of exposure to infective bites by mosquitoes.^46^ Thus, in high transmission areas, not all malaria infections cause symptoms as some individuals may remain asymptomatic even when an individual have high parasitaemia.^47^ When such individuals develop symptoms, they will test positive for malaria while malaria may potentially not be the direct cause of the symptoms.^13^ As a result, any individual who tests positive for malaria by RDT will be considered not to have NMFIs (based on the criteria used in this study) resulting in a low prevalence of NMFIs. On the contrary, in areas with low malaria burden, most of the febrile patients will test negative for malaria thus reducing the masking effect of malaria infections and resulting in high NMFI prevalence. Therefore, while it may be true that in some areas where the prevalence of NMFIs is high and the burden of malaria is low, co-infections with malaria which is common in areas of high transmission intensities may introduce bias and act as a confounder. Thus, other causes of fever should always be investigated even in malaria-positive individuals, particularly in high malaria-burden regions although this may be challenging under clinical conditions.^13^

This study showed high variations in NMFI prevalence among patients with different age groups, with adults exhibiting the highest prevalence than under-fives and school children. The increased prevalence in adults suggests that the adult population may be more exposed to febrile illnesses due to increased mobility, occupational exposure, and sometimes immunosenescence. Taking more responsibility, adults may be exposed to multiple and higher doses of infectious agents than younger children. Increased exposure frequently raises the chance of contracting an infection, but there is no apparent evidence linking this risk to the findings of this study.^48^ The observed higher prevalence in under-fives could be due to the vulnerability of this group to infections. These findings are similar to those reported in Malawi where 72.0% of under-fives and 77.0% of adults had NMFIs.^44,49,50^ The low prevalence of NMFIs among school children could be attributed to the high prevalence of malaria in this group as previously reported by other studies,^44,49,50^ since febrile patients were excluded from NMFI analysis if they tested positive for malaria by RDT. Additionally, the low prevalence of NMFI among school children could be attributed to their relatively stronger immunity acquired through repeated antigenic exposure. It may also be because, at the age of 5 - 15, the immune system is just starting to mature. As a result, most infections can be prevented by the immune system, making this group less susceptible to most NMFI-causing pathogens.^51,52^ This is consistent with previous research conducted elsewhere, where school children had the lowest prevalence of NMFI compared to under-fives and adults.^44,49^ The pattern observed in this study is similar to what was reported by other studies on malaria-related fevers, where school children typically showed a higher prevalence making them have NMFIs in the absence of tests for other pathogens.^44,49^ This results in a low prevalence of non-malarial fevers since a large proportion of the fevers is parasitologically attributed to malaria even if there is no evidence of clinical association unless multiple tests are done to rule out any other infections by fever-causing pathogens.

The slightly higher NMFI prevalence observed in females compared to males may be attributed to a variety of social, biological, and healthcare-seeking behaviours. In many settings, women are more likely to seek healthcare services, leading to higher detection rates of febrile illnesses. Additionally, biological differences such as immune response variations between sexes may also contribute to this observation.^44,49,50^ Due to social and biological factors, the prevalence of malaria is usually high in males compared to females,^44,49,50^ which results in the exclusion of more males than females in the NMFI analysis, since the potential for malaria co-infection with other febrile aetiologies was not pursued in this study. However, further research is needed to fully understand the underlying causes of this disparity. The findings of higher odds of NMFIs are consistent with what was reported by other studies which showed a higher prevalence of NMFIs in females (70.0%) compared to males (68.0%).^29^

The key strength of the study is its extensive coverage, utilising a large number of samples from all 26 regions of Mainland Tanzania, thus providing a comprehensive representation of the country’s diverse population and transmission settings. However, this study had some limitations. Being a cross-sectional study, the data may not be a true representation of the actual prevalence of NMFIs in a given area because the study only captured a single time point and, hence had a limited temporal scope. Also, the data for this study were collected as part of the MSMT project whose sites were purposively selected to maximise the recruitment of patients with malaria based on the detection of RDT-positive results. Therefore, the sites may be biased towards high transmission areas even in low transmission regions given the current heterogeneous nature of malaria in Mainland Tanzania^56,57^. Thus, future studies should consider inclusive sampling covering areas of different levels of malaria burden, preferably using DHIS2 data for a more comprehensive mapping of NMFIs country-wide. Additionally, this study could not explore the specific causes of NMFIs because it was not designed to offer POC or laboratory tests for other pathogens. However, it has highlighted the magnitude of this problem in the entire country, with a higher burden of NMFIs particularly in areas of low transmission intensities as previously reported by other studies.^13^ Analysis to address this issue will need to be performed and reported in future studies to appropriately inform policy and decisions for appropriately targeting and better case management and the control of NMFIs.

## Conclusion

Non-malarial febrile illnesses were fairly common and their burden and distribution patterns varied significantly across regions and malaria transmission strata. Females, under-fives and individuals from areas with low and very low transmission intensities of malaria had a higher prevalence of NMFIs and an increased likelihood of experiencing non-malarial fevers compared to males, adults, school children and individuals living in settings with high malaria transmission intensities. These findings have important implications for healthcare planning, offering critical insights to enhance the diagnosis and management of febrile illnesses beyond malaria. The results further suggest that the identified groups of patients should be appropriately targeted with POC tests and treatment strategies. By country-wide mapping of NMFIs, this research contributes to a better understanding of the local epidemiology of NMFIs and supports the development of more effective management and control strategies. Addressing the gaps in evidence-based information on NMFIs will ultimately lead to improved patient outcomes and better healthcare delivery in regions with varying endemicity of malaria and other fever-causing pathogens.

## Data Availability

The data used in this paper are available and can be obtained upon reasonable request from the corresponding author institutional approval by NIMR, and the signing of a data transfer agreement between the donor and recipient.

## Acknowledgements

The authors wish to thank all patients and parents/guardians of all the children who participated in the survey. We acknowledge the contributions of the following project staff and other colleagues who participated in the data collection: Hussein Semboja, Sharifa Hassan, Ezekiel Malecela, Godwin Gaudin, Ildephonce Mathias, Kusa Mchaina, Oswald Oscar, and Tilaus Gustav. Many thanks to the finance, administrative and logistic support team at NIMR: Christopher Masaka, Millen Meena, Beatrice Mwampeta, Gracia Sanga, Neema Manumbu, Arison Ekoni, Twalipo Mponzi, Alfred Sezary, Sadiki Yusuph, and Rodrick Ulomi. The authors also extend their gratitude to the management of NIMR, NMCP and PO-RALG (including the regional administrative secretaries of the 26 regions and district officials), and staff from all 86 health facilities. Authors received and appreciate the technical and logistics support from partners at Brown University, the University of North Carolina at Chapel Hill, the CDC Foundation and the Bill and Melinda Gates Foundation team.

## Disclosures regarding real or perceived conflicts of interest

The authors declare that they have no competing interests.

## Author’s current addresses

Salehe S. Mandai, National Institute for Medical Research, Dar es Salaam, Tanzania

Angelina J. Kisambale, National Institute for Medical Research, Dar es Salaam, Tanzania

Daniel A. Petro, University of Dar es Salaam, Dar es Salaam, Tanzania

Catherine Bakari, National Institute for Medical Research, Dar es Salaam, Tanzania

Gervas A. Chacha, National Institute for Medical Research, Dar es Salaam, Tanzania

Rule Bododo, National Institute for Medical Research, Dar es Salaam, Tanzania

Rashid A. Madebe, National Institute for Medical Research, Dar es Salaam, Tanzania

Dativa Pereus, National Institute for Medical Research, Dar es Salaam, Tanzania

Daniel P. Challe, National Institute for Medical Research, Tanga Research Center, Tanga, Tanzania

Ramadhani Moshi, National Institute for Medical Research, Dar es Salaam, Tanzania

Ruth A. Mbwambo, National Institute for Medical Research, Dar es Salaam, Tanzania

Grace Kiiza Kanyankole, National Institute for Medical Research, Dar es Salaam, Tanzania

Sijenunu Aaron, National Malaria Control Programme, Dodoma, Tanzania

Daniel Mbwambo, National Malaria Control Programme, Dodoma, Tanzania

Stella Kajange, President’s Office-Regional Administrative Local Government, Dodoma Tanzania

Samwel Lazaro, National Malaria Control Programme, Dodoma, Tanzania

Ntuli Kapologwe, Directorate of Preventive Services, Ministry of Health, Dodoma, Tanzania

Celine I. Mandara, National Institute for Medical Research, Dar es Salaam, Tanzania

Misago D. Seth, National Institute for Medical Research, Dar es Salaam, Tanzania

Deus S. Ishengoma, National Institute for Medical Research, Dar es Salaam, Tanzania

