## Supplementary Table 1 for "Mapping of the country-wide prevalence of non-malarial febrile illnesses in areas with varying malaria transmission intensities in Mainland Tanzania"

**Supplementary Table 1: Prevalence of non-malaria febrile illnesses (NMFIs) by region, sex, and age group.**

| **Regions** | | | | **Sex** | | | | | | **Age group** | | | | | | | | |
| --- | --- | --- | --- | --- | --- | --- | --- | --- | --- | --- | --- | --- | --- | --- | --- | --- | --- | --- |
|  |  |  |  | **Female** | | | **Male** | | | **< 5 years** | | | **5 -< 15 years** | | | **≤ 15 years** | | |
|  | Tested for malaria | Negative malaria tests | Proportion of NMFI (%) | Tested for malaria | Negative malaria tests | Proportion of NMFI (%) | Tested for malaria | Negative malaria tests | Proportion of NMFI (%) | Tested for malaria | Negative malaria tests | Proportion of NMFI (%) | Tested for malaria | Negative malaria tests | Proportion of NMFI (%) | Tested for malaria | Negative malaria tests | Proportion of NMFI (%) |
| Arusha | 561 | 263 | 46.9 | 302 | 166 | 55.0 | 259 | 97 | 37.5 | 198 | 105 | 53.0 | 94 | 27 | 28.7 | 269 | 131 | 48.7 |
| Dar es Salaam | 1117 | 792 | 70.9 | 559 | 412 | 73.7 | 558 | 380 | 68.1 | 340 | 286 | 84.1 | 161 | 108 | 67.1 | 616 | 398 | 64.6 |
| Dodoma | 841 | 514 | 61.1 | 482 | 312 | 64.7 | 359 | 202 | 56.3 | 320 | 209 | 65.3 | 138 | 77 | 55.8 | 383 | 228 | 59.5 |
| Geita | 461 | 131 | 28.4 | 261 | 82 | 31.4 | 200 | 49 | 24.5 | 245 | 82 | 33.5 | 95 | 13 | 13.7 | 121 | 36 | 29.8 |
| Iringa | 95 | 58 | 61.1 | 47 | 27 | 57.4 | 48 | 31 | 64.6 | 44 | 31 | 70.5 | 26 | 10 | 38.5 | 25 | 17 | 68.0 |
| Kagera | 2352 | 965 | 41.0 | 1360 | 541 | 39.8 | 992 | 424 | 42.7 | 1333 | 549 | 41.2 | 328 | 130 | 39.6 | 691 | 286 | 41.4 |
| Katavi | 578 | 248 | 42.9 | 377 | 159 | 42.2 | 201 | 89 | 44.3 | 201 | 103 | 51.2 | 107 | 38 | 35.5 | 270 | 107 | 39.6 |
| Kigoma | 460 | 112 | 24.3 | 261 | 73 | 28.0 | 199 | 39 | 19.6 | 240 | 50 | 20.8 | 60 | 9 | 15.0 | 160 | 53 | 33.1 |
| Kilimanjaro | 2154 | 1844 | 85.6 | 1272 | 1135 | 89.2 | 882 | 709 | 80.4 | 308 | 281 | 91.2 | 238 | 193 | 81.1 | 1608 | 1370 | 85.2 |
| Lindi | 426 | 96 | 22.5 | 251 | 62 | 24.7 | 175 | 34 | 19.4 | 269 | 59 | 21.9 | 49 | 6 | 12.2 | 108 | 31 | 28.7 |
| Manyara | 596 | 261 | 43.8 | 335 | 153 | 45.7 | 261 | 108 | 41.4 | 165 | 88 | 53.3 | 113 | 43 | 38.1 | 318 | 130 | 40.9 |
| Mara | 907 | 279 | 30.8 | 525 | 153 | 29.1 | 382 | 126 | 33.0 | 246 | 106 | 43.1 | 270 | 66 | 24.4 | 391 | 107 | 27.4 |
| Mbeya | 560 | 243 | 43.4 | 332 | 144 | 43.4 | 228 | 99 | 43.4 | 208 | 95 | 45.7 | 82 | 30 | 36.6 | 270 | 118 | 43.7 |
| Morogoro | 476 | 146 | 30.7 | 315 | 107 | 34.0 | 161 | 39 | 24.2 | 184 | 47 | 25.5 | 98 | 21 | 21.4 | 194 | 78 | 40.2 |
| Mtwara | 820 | 196 | 23.9 | 472 | 116 | 24.6 | 348 | 80 | 23.0 | 522 | 119 | 22.8 | 78 | 13 | 16.7 | 220 | 64 | 29.1 |
| Mwanza | 455 | 113 | 24.8 | 251 | 69 | 27.5 | 204 | 44 | 21.6 | 222 | 64 | 28.8 | 106 | 12 | 11.3 | 127 | 37 | 29.1 |
| Njombe | 402 | 62 | 15.4 | 225 | 42 | 18.7 | 177 | 20 | 11.3 | 102 | 14 | 13.7 | 104 | 16 | 15.4 | 196 | 32 | 16.3 |
| Pwani | 667 | 332 | 49.8 | 380 | 193 | 50.8 | 287 | 139 | 48.4 | 315 | 163 | 51.7 | 121 | 42 | 34.7 | 231 | 127 | 55.0 |
| Rukwa | 662 | 274 | 41.4 | 397 | 166 | 41.8 | 265 | 108 | 40.8 | 267 | 117 | 43.8 | 150 | 46 | 30.7 | 245 | 111 | 45.3 |
| Ruvuma | 526 | 194 | 36.9 | 280 | 113 | 40.4 | 246 | 81 | 32.9 | 245 | 93 | 38.0 | 122 | 32 | 26.2 | 159 | 69 | 43.4 |
| Shinyanga | 430 | 84 | 19.5 | 250 | 49 | 19.6 | 180 | 35 | 19.4 | 219 | 54 | 24.7 | 97 | 8 | 8.2 | 114 | 22 | 19.3 |
| Simiyu | 477 | 140 | 29.4 | 306 | 102 | 33.3 | 171 | 38 | 22.2 | 250 | 95 | 38.0 | 74 | 5 | 6.8 | 153 | 40 | 26.1 |
| Singida | 524 | 179 | 34.2 | 298 | 108 | 36.2 | 226 | 71 | 31.4 | 341 | 118 | 34.6 | 51 | 14 | 27.5 | 132 | 47 | 35.6 |
| Songwe | 464 | 147 | 31.7 | 269 | 87 | 32.3 | 195 | 60 | 30.8 | 156 | 51 | 32.7 | 118 | 28 | 23.7 | 190 | 68 | 35.8 |
| Tabora | 1004 | 393 | 39.1 | 569 | 216 | 38.0 | 435 | 177 | 40.7 | 605 | 253 | 41.8 | 101 | 35 | 34.7 | 298 | 105 | 35.2 |
| Tanga | 553 | 207 | 37.4 | 348 | 126 | 36.2 | 205 | 81 | 39.5 | 139 | 44 | 31.7 | 141 | 48 | 34.0 | 273 | 115 | 42.1 |
| **Total** | **18568** | **8273** | **44.6** | **10724** | **4913** | **45.8** | **7844** | **3360** | **42.8** | **7684** | **3276** | **42.6** | **3122** | **1070** | **34.3** | **7762** | **3927** | **50.6** |
